# Structural and functional MRI changes associated with cognitive impairment and dementia in Parkinson disease

**DOI:** 10.1101/19002147

**Authors:** Conor Owens-Walton, David Jakabek, Brian D. Power, Mark Walterfang, Sara Hall, Danielle van Westen, Jeffrey C.L. Looi, Marnie Shaw, Oskar Hansson

## Abstract

**Objective:** Cognitive impairment in Parkinson disease (PD) places a high burden on patients and the pathophysiological mechanisms that differentiate patients with cognitive impairment from those without it are still incompletely understood.

**Methods:** We conducted a resting-state seed region-of-interest approach to investigate functional connectivity (FC) of important subdivisions of the caudate nucleus, putamen and thalamus in controls (n=33), cognitively unimpaired subjects (PD-CU, n=33), PD subjects with mild cognitive impairment (PD-MCI, n=22) and subjects with dementia (PDD, n=17). We then investigated how the morphology of these structures differed between groups.

**Results:** PD-CU subjects, compared to controls, displayed increased and decreased FC of caudate, putamen and thalamic subdivisions. PD-MCI subjects demonstrated reduced FC of the mediodorsal thalamus with the paracingulate cortex, compared to PD-CU, and reduced FC of the mediodorsal thalamus with the posterior cingulate cortex compared to PDD. Extensive volumetric and surface-based deflation was found in subjects with dementia.

**Conclusions:** Our research suggests that cognitive impairment and dementia in PD may be associated with a breakdown in FC of the mediodorsal thalamus with para- and posterior cingulate regions of the brain, respectively.

**Significance:** Cognitive impairment and dementia in PD may relate to pathophysiological changes in FC of the mediodorsal thalamus.

## 1. Introduction

Parkinson disease (PD) is the second most common neurodegenerative disorder in the world, affecting 2-3% of the population over the age of 65 (Poewe et al., 2017). Traditionally categorized as a disorder of movement, it is now widely recognised that patients experience significant neuropsychiatric symptoms relating to executive, memory, attentional and visual disturbances (Litvan et al., 2012). Recent recognition of the high prevalence and substantial impact of cognitive impairment in PD has drawn research attention to PD-related mild cognitive impairment (MCI)^1^ (Goldman et al., 2018) which is associated with a significantly higher likelihood of developing dementia (Broeders et al., 2013).

The pathophysiological mechanisms that distinguish PD patients with cognitive impairment from those without it are poorly understood (Williams-Gray et al., 2007). The pathological hallmark of the disorder is the presence of α-synuclein-immunopositive Lewy bodies and neurites (Obeso et al., 2000) which results in a loss of dopaminergic neurons in the substantia nigra pars compacta, and subsequent depletion of dopamine at the striatum (caudate nucleus and putamen) (Kish et al., 1988). While nigrostriatal dopamine loss is a core feature of PD, evidence suggests that pathology within the thalamus also contributes to the abnormal neural activity associated with the disorder (Halliday, 2009). The combined neural activity in these nodes within basal ganglia-thalamocortical circuits in PD results in brain network abnormalities being as key components to PD pathophysiology (Strafella, 2013).

To understand the functioning of brain networks it is necessary to study both the constituent neuronal elements of networks, as well as their interconnections (Sporns et al., 2005). Investigating neuronal elements can be done by quantifying disease-related effects on the morphology of ‘hubs’ (Looi et al., 2014) while interconnections can be investigated via resting-state functional connectivity (FC) analyses (Damoiseaux et al., 2006). Due to the importance of the caudate nucleus, putamen and the thalamus to the abnormal neural activity associated with PD, coupled with the fact that these structures are considered important hubs in brain networks (Hwang et al., 2017, Looi and Walterfang, 2013), an analysis of the morphology and FC of these structures may yield neuroimaging biomarkers of brain network abnormalities in PD that relate to cognitive dysfunction.

There is a significant variability in the field of neuroimaging biomarkers in PD, with reviews of structural and functional studies revealing few clear patterns (Khan et al., 2018). Research has yielded important information about the functioning of widescale intrinsic FC networks is impacted in PD relative to cognitive status, however the specific role played by important hubs within basal ganglia-thalamocortical circuitry is yet to be fully elucidated. Studies have found cognitively unimpaired PD is associated with both increases (Gorges et al., 2015) and decreases (Bell et al., 2015) in FC of basal ganglia-thalamocortical circuitry, while PD subjects with MCI have been shown to generally display decreases in FC (Amboni et al., 2015, Gorges et al., 2015). Data on the morphological changes to the caudate, putamen and thalamus in PD subjects without dementia is also varied. Research groups have demonstrated both the presence (Garg et al., 2015, Mckeown et al., 2008) and absence (Menke et al., 2014, Messina et al., 2011) of morphological changes to the thalamus, while others have shown either the presence (Pitcher et al., 2012, Sterling et al., 2013) or absence (Menke et al., 2014, Owens-Walton et al., 2018) of morphological changes to the caudate or putamen. Studies suggest that PD with dementia is associated with atrophic changes to the caudate nucleus, putamen and thalamus (Summerfield et al., 2005, Owens-Walton et al., 2018), however the presence and extent of such changes in PD-MCI is uncertain (Chen et al., 2016, Melzer et al., 2012). Using a well-defined clinical cohort, and a targeted resting-state fMRI FC approach, the current study aims to better understand how FC and morphology of the caudate nucleus, putamen and thalamus are impacted in PD, and how these factors may vary between cognitively unimpaired PD patients, PD-MCI and PD patients with dementia.

## 2. Methods

### 2.1 Participants

Participants in this study were from the Swedish BioFINDER study (www.biofinder.se) and gave informed written consent. All elements of the research were performed in accordance with the World Medical Association’s Declaration of Helsinki and ethical approval was obtained through the Ethical Review Board of Lund, Sweden, and the Human Research Ethics Committee at the Australian National University, Canberra, Australia.

### 2.2 PD disease groups: cognitive impairment

Diagnosis of PD was made by a neurologist using the National Institute of Neurological and Stroke Diagnostic Criteria (Gelb et al., 1999). The total PD cohort was subdivided into three cognitive impairment disease groups. The first group was made up of cognitively unimpaired PD patients (PD-CU; *n* = 33) while the second group was made up of PD patients with mild cognitive impairment (PD-MCI; *n* = 22), based on the Movement Disorder Society Task force guidelines (Litvan et al., 2012). The MCI criterion is met when patients score at least 1 standard deviation below the normative mean in at least two cognitive tests, including executive function, attention, visuospatial, memory and language domains. The third subgroup consists of PD participants with dementia (PDD; *n* = 17) diagnosed using the Clinical Diagnostic Criteria for Dementia Associated with PD (Emre et al., 2007). A healthy control group (Controls) was included for comparative purposes (*n* = 26). Exclusion criteria for this study included refusing MRI, the presence of significant alcohol or substance misuse and/or a significant unstably systematic illness or organ failure. All participants underwent a cognitive and neurological examination by a medical doctor with extensive experience with movement disorders, while PD patients remained on their usual medication regimes. Clinical functioning of participants in the current study is demonstrated by performance on the Hoehn and Yahr, assessing the progression of the disorder, the ‘Unified Parkinson’s Disease Rating Scale Part-III’ test (UPDRS-III), assessing the motor signs of PD (Fahn and Elton, 1987), and the ‘Mini Mental State Examination’ (MMSE), assessing cognitive mental state (Folstein et al., 1975).

### 2.3 MRI acquisition

Structural MRI data was obtained on a 3T scanner (Trio, Siemens Magnetom, Erlangen, Germany) with a 20-channel head-coil. High resolution T1-weighted three-dimensional anatomical brain images were acquired using a magnetization-prepared rapid acquisition technique with gradient-echo sequence (repetition time = 7 ms; echo time = 3 ms; flip angle = 90 degrees; voxel size = isotropic 1mm^3^). Image matrix size was 356 voxels in the coronal and sagittal planes and 176 voxels in the axial plane. Functional MRI data consists of 256 T2*-weighted echo planar imaging volumes (repetition time = 1850 ms; echo time 30 ms; flip angle = 90 degrees; voxel size 3 × 3 × 3.75 mm^3^) for each participant. Matrix size was 64 voxels in the coronal and sagittal planes and 36 voxels in the axial plane. Subjects were instructed to lie still with their eyes closed, not to fall asleep and not to think of anything in particular during the scan, which lasted for approximately 8 minutes.

### 2.4 Preprocessing of T1 anatomical MRI data

Preprocessing of T1-weighted structural data was performed using the FSL software package (FSL, Oxford, UK; version 5.0.10) (Jenkinson et al., 2012). Images underwent brain extraction using the FSL-BET tool (Smith, 2002) which were then processed using FSL-FAST, segmenting the T1-image into brain tissue types whilst correcting for image intensity fluctuations caused by inhomogeneities in the radio frequency field (Zhang et al., 2001).

### 2.5 Preprocessing of resting-state fMRI data

Preprocessing of resting-state functional images was performed using FSL-FEAT (v6.0). Steps included deletion of first 6 volumes, motion correction using FSL-MCFLIRT (Jenkinson et al., 2002) and removal of non-brain tissue using FSL-BET (Smith, 2002). Registration of functional images to subject-specific high resolution T1-weighted structural images using a boundary-based registration (Greve and Fischl, 2009) within FSL-FLIRT linear registration tool (Jenkinson et al., 2002, Jenkinson and Smith, 2001). Registration of functional images to MNI 152 T1 2mm^3^ standard space was performed with FSL-FLIRT with 12 degrees of freedom, further refined using FSL-FNIRT nonlinear registration (Andersson et al., 2007a, Andersson et al., 2007b) with a warp resolution of 10mm and a resampling resolution of 4mm. Data denoising was then performed with FSL-MELODIC (v3.15) independent component analysis (Beckmann and Smith, 2004) to identify any potential influence of head motion, scanner and cerebrospinal fluid artefacts. FSL-FIX (v1.06) (Salimi-Khorshidi et al., 2014) was then used to remove components classified as ‘noise’ and also correct for motion confounds. Spatial smoothing was then performed (FWHM Gaussian kernel = 5mm) followed by a multiplicative mean intensity normalization and a high-pass temporal filtering Gaussian-weighted least-squares straight line fitting with sigma = 50.0s (Woolrich et al., 2001). For a full explanation of the denoising procedure see ‘1. Supplementary Information Materials and Methods: *1.1 Independent component analysis denoising*.’

### 2.6 Resting-state fMRI: Seed-based region-of-interest approach

To investigate the role of the caudate, putamen and thalamus in PD groups with varying levels of cognitive impairment, we used an atlas-based method to subdivide the structures into functionally relevant regions-of-interest (ROI). We performed this step using the Oxford Thalamic Connectivity Atlas (Behrens et al., 2003) and the Oxford-GSK-Imanova Connectivity Atlas (Tziortzi et al., 2014) within FSLEyes. Regions of the dorsal and central caudate nucleus (Figure 1) and the anterior putamen have the highest probability of connectivity with the anterior and dorsolateral prefrontal cortices in the brain (BA9, 9/46 and 10) and are said to be involved in cognitive processes, including perception, memory, reasoning and judgement (Haber, 2003). The mediodorsal and anterior thalamic nuclei have the highest probability of connectivity to the dorsolateral and prefrontal cortices (Behrens et al., 2003) and these thalamic nuclear regions are said to be involved in limbic, episodic memory and cognitive functions (Power and Looi, 2015). For ease of reference, we will refer to these three seed-ROIs as the ‘dorsal caudate’, ‘anterior putamen’ and ‘mediodorsal thalamus’ respectively. These three bilateral standard space MNI 152 2mm^3^ seed-ROI masks were thresholded to only include voxels that had a greater than 50% likelihood of inclusion within the mask (Figure 1A). Masks were then registered to subject-specific structural T1 space where we used them to ‘cookie-cut’ ROIs from the FSL-FIRST segmentation masks. This was done by overlaying the seed mask over the FSL-FIRST segmentation mask, with the seed-ROI being the neuroanatomical area covered by both masks. The created seed-ROIs were then converted to subject-specific functional MRI space where we performed a final erosion to avoid partial volume effects (Caballero-Gaudes and Reynolds, 2017). Mean activation data from within the three seed-ROIs at each timepoint was used as explanatory variables for subsequent statistical analyses. Before investigating the differences in FC between groups we conducted three one-sample *t*-tests on all of the Control subjects to look at the FC of our seed-ROIs with the rest of the brain. This step indicates the areas of highest FC with our seed-ROIs and provides a backdrop on which to interpret our results (Figure 1B).

**Figure 1:**
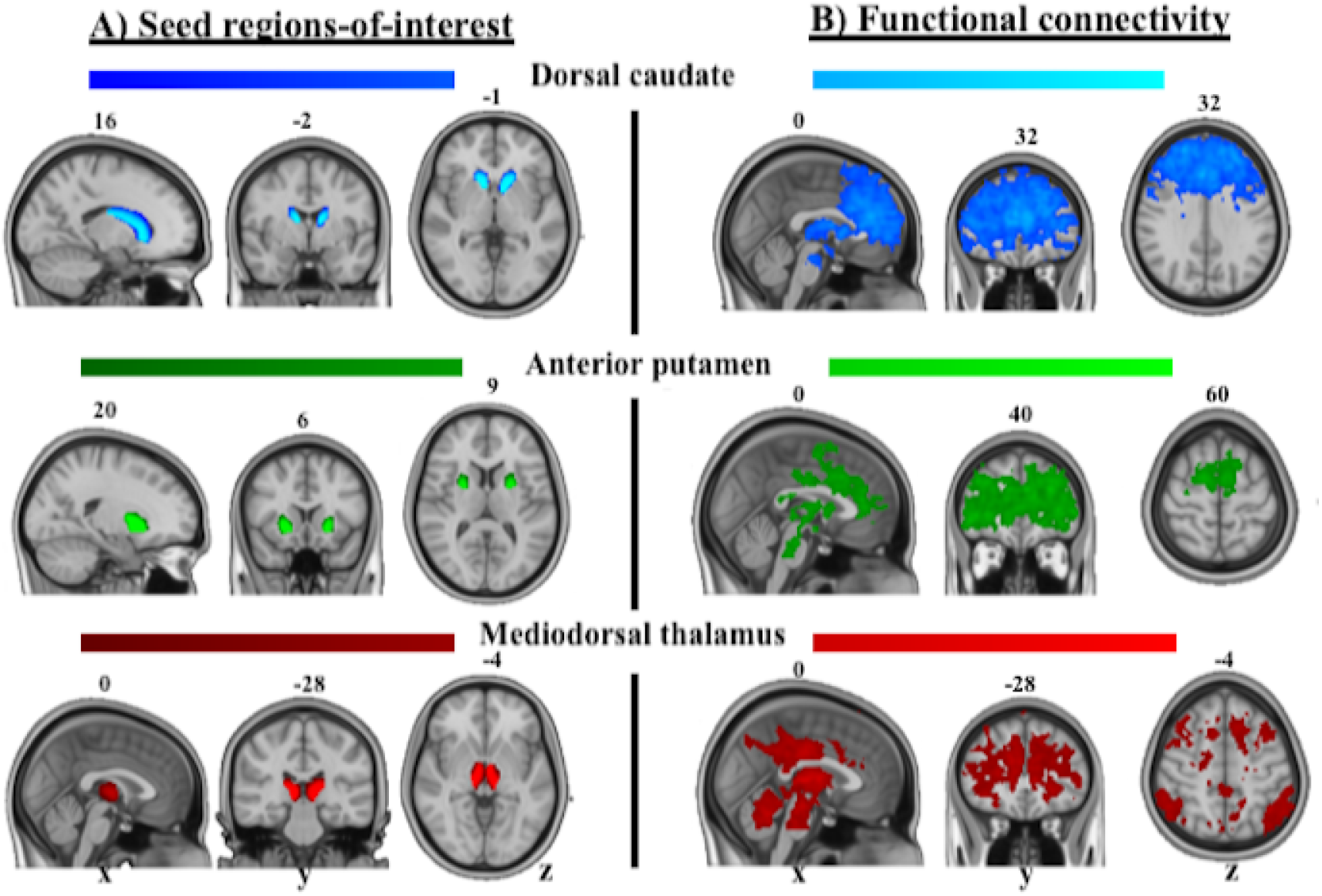
Seed regions-of-interest and one-sample *t*-tests demonstrating putative functional connectivity pathways in Control subjects. A) Subject specific seed regions-of-interest masks were converted to MNI 152 T1 standard space and combined to produce this image. Lighter areas of each colour are indicative of a greater proportion of voxels in that area being included in the relevant mask. B) Z-statistic (Gaussianized) images from Control participant’s individual-level GLM statistical maps, thresholded non-parametrically using clusters determined by Z > 3.1 and a (corrected) cluster significance of *p* < 0.05.

### 2.7 Resting-state fMRI: Statistical analyses

Three individual-level FC analyses were performed for each participant using a general linear model (GLM) in a mass univariate voxelwise whole-brain analysis (Woolrich et al., 2004). This GLM approach measures the correlation between activity within each of the three seed-ROIs and the rest of the brain. We used a FILM pre-whitening correction (Woolrich et al., 2001) and shifted the model with a temporal derivative to account for slice timing effects and variations in the hemodynamic response function. As there were significant differences in UPDRS-III motor scores, we performed extra processing steps to minimise possible confounding effects of head motion, as well as regressing out signals from within white matter, ventricle and whole-brain masks. For a full explanation these steps, please see ‘1. Supplementary Information Materials and Methods: *1.2 Individual-level GLM functional connectivity analyses*.’ Group-level analyses were performed using FSL-FLAME in a mixed-effects design (Woolrich et al., 2004). Age and gender data were included as covariates. When investigating FC between the PD-CU, PD-MCI and PDD groups we also included levodopa-equivalent daily dosage (LEDD) data as a covariate. Z-statistic (Gaussianized) images from participants individual-level GLM statistical maps were thresholded non-parametrically using clusters determined by Z > 3.1 and a (corrected) cluster significance of *p* < 0.05 (Worsley, 2001).

### 2.8 Volumetrics: Statistical analyses

Volumes of the caudate, putamen and thalamus were estimated using the segmentation and registration tool FSL-FIRST (Patenaude et al., 2011). This uses a model-based approach to segment subcortical structures producing output meshes and volumetric data. Investigating differences between experimental groups was performed with SPSS 22.0 (IBM Corporation, Somers, New York, USA) utilising two-tailed multivariate analysis of covariance models controlling for intracranial volume (derived from FSL-FAST) and age. Multivariate partial eta squared values (η^2^) are used to report effect sizes (Cohen, 1992).

### 2.9 SPHARM-PDM surface shape: Statistical analyses

Volumetric analyses can investigate overall atrophic differences between groups, however they cannot highlight *where* this atrophy takes place. Accordingly, we used spherical harmonic parametrization three-dimensional point distribution model (SPHARM-PDM) to investigate localized surface changes to the segmentations of the caudate, putamen and thalamus between experimental groups (Styner et al., 2006). This method produces mean displacement maps which show the magnitude of surface displacement (deflation or inflation in mm) between corresponding points on the mean surface of one experimental group compared to another. Segmentations were preprocessed to ensure a spherical topology, then described by spherical harmonic functions and sampled onto surfaces of 1002 points. Surfaces were then aligned using a rigid-body Procrustes alignment to a mean template created from a group sample. Comparisons between groups were performed using multivariate analysis of covariance models with a Hotelling statistic in R (version 3.2.1, R Development Core Team, 2014). All analyses included covariates of estimated total intracranial volume, age and sex. Significant surface changes are indicated by regions with a *p* value < 0.05, corrected for multiple comparisons with a false-discovery rate (FDR) bound *q* of 5% (Genovese et al., 2002). Results are presented as a composite map of each structure with displacement only displayed in regions that are FDR significant.

## 3. Results

### 3.1 Participant characteristics

The characteristics of participants are displayed in Table 1. A chi-square test for independence found no significant difference in sex between the groups [χ^2^ (3, *n* = 98) = 0.89, *p* = 0.86]. One-way analyses of variance found no significant difference in age [*F* (3, 94) = 2.169, *p* = 0.10], ICV [*F* (3, 94) = 0.323, *p* = 0.81], years of education [*F* (3, 79) = 1.255, *p* = 0.30] or average DVARS [F (3, 94) = 2.460, *p* = 0.07] between groups (Table 1).

**Table 1:** Participant characteristics.

| Item | Controls | PD-CU | PD-MCI | PDD | p-value |
| --- | --- | --- | --- | --- | --- |
| Number of participants | 26 | 33 | 22 | 17 | - |
| Female/Male | 12/14 | 15/18 | 12/10 | 7/10 | 0.86 |
| Average age in years | 69.59<br>$\pm 6.17$ | 69.25<br>$\pm 6.31$ | 70.53<br>$\pm 5.38$ | 73.70<br>$\pm 6.68$ | 0.10 |
| ICV (cm<sup>3</sup>) | 1456.00<br>$\pm 135.93$ | 1491.68<br>$\pm 139.08$ | 1471.06<br>$\pm 132.13$ | 1481.80<br>$\pm 168.95$ | 0.81 |
| Years of education | 12.73<br>$\pm 3.41$ | 11.34<br>$\pm 5.36$ | 9.66<br>$\pm 6.81$ | 11.50<br>$\pm 4.52$ | 0.30 |
| DVARS | 11.97<br>$\pm 3.21$ | 14.96<br>$\pm 3.87$ | 17.46<br>$\pm 12.71$ | 15.62<br>$\pm 6.39$ | 0.07 |
| Years since diagnosis | - | 6.30<br>$\pm 5.24$ | 5.54<br>$\pm 3.56$ | 11.88<br>$\pm 5.48$ | - |
| LEDD | - | 639.60<br>$\pm 485.23$ | 610.73<br>$\pm 262.11$ | 1010.72<br>$\pm 625.53$ | - |
| H&Y | - | 1.742<br>$\pm 0.73$ | 2.00<br>$\pm 0.72$ | 2.853<br>$\pm 0.77$ | - |
| UPDRS-III | 2.35<br>$\pm 2.84$ | 12.61<br>$\pm 9.55$ | 14.27<br>$\pm 9.07$ | 33.00<br>$\pm 10.21$ | <0.01 |
| MMSE | 28.69<br>$\pm 1.16$ | 28.76<br>$\pm 1.00$ | 27.41<br>$\pm 1.65$ | 22.88<br>$\pm 3.77$ | <0.01 |

**Key:** Participant data presented as mean ± standard deviation. All data corrected to two decimal places. Abbreviations: ICV, total intracranial volume; DVARS, average image intensity of each frame $n$ in relation to $n+1$ ; LEDD, levodopa-equivalent daily dosage (mg); H&Y, Hoehn and Yahr Scale; UPDRS-III, Unified Parkinson's Disease Rating Scale, part-III; MMSE, Mini Mental-state Examination.

### 3.2 Resting-state seed-based functional connectivity analyses

#### 3.2.1 PD-CU compared to Controls

##### Dorsal caudate

Analysis of the dorsal caudate in PD-CU compared to Controls found clusters of increased FC with the right superior frontal gyrus (BA8), left frontal pole (BA10) and right middle frontal gyrus (BA8) (Figure 2A, Table 2). These clusters extended to include local maxima at the left postcentral gyrus (BA3), left precentral gyrus (BA6), left superior frontal gyrus (BA6), left paracingulate gyrus (BA32), right frontal pole (BA10), left anterior cingulate (BA24) and the left middle frontal gyrus (BA8) (Figure 2A; Supplementary Information Table 1). We found clusters of decreased FC at the right lateral occipital cortex (BA19), left cerebellar crus I, left angular gyrus (BA39) and left cerebellar lobule VI (Figure 2A; Table 2). These clusters extended to include local maxima at the left supramarginal gyrus (BA39) and left cerebellar vermis VI (Figure 2A; Supplementary Information Table 1).

**Table 2:**
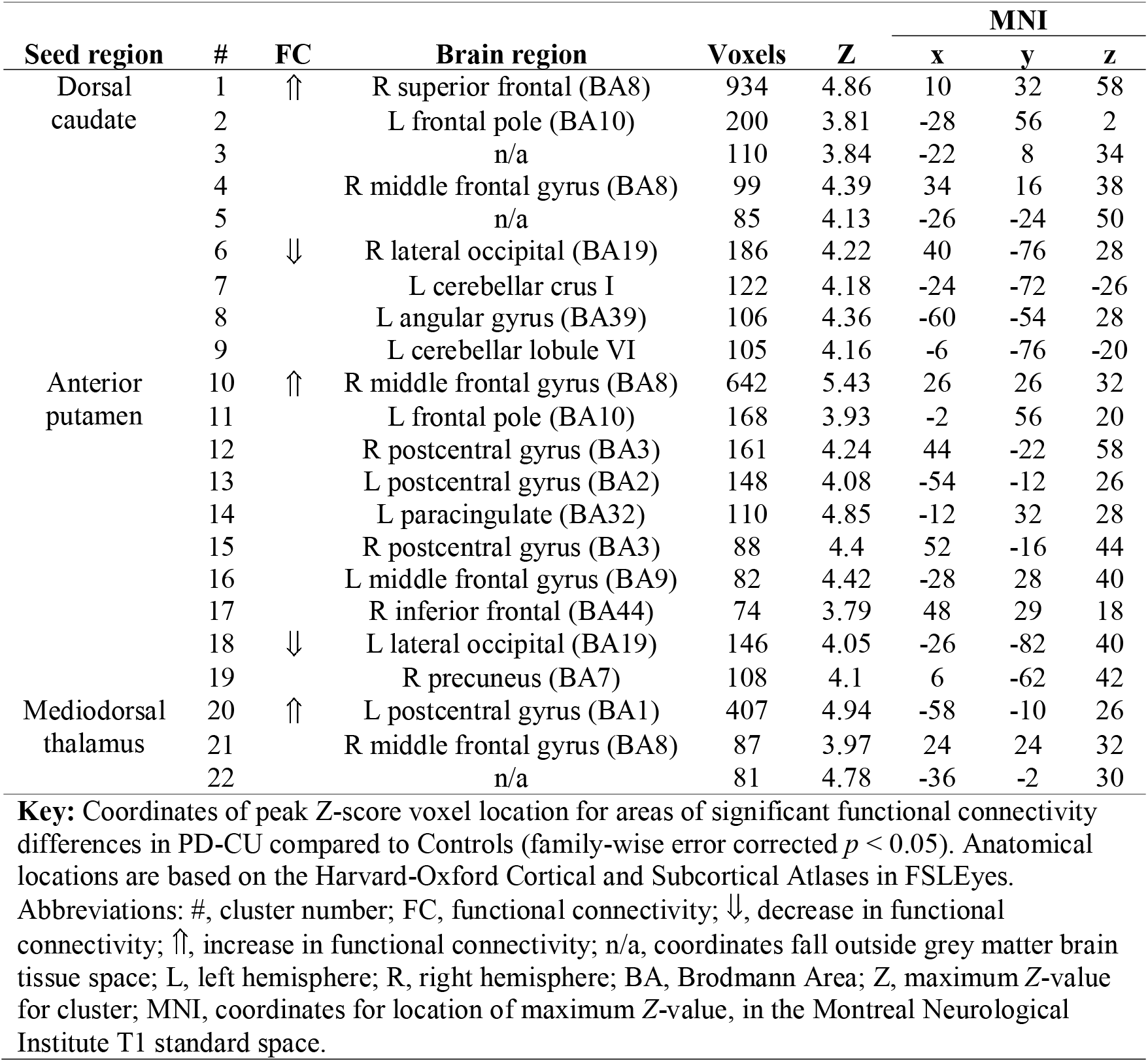
Functional connectivity differences in PD-CU compared to Controls.

**Figure 2:**
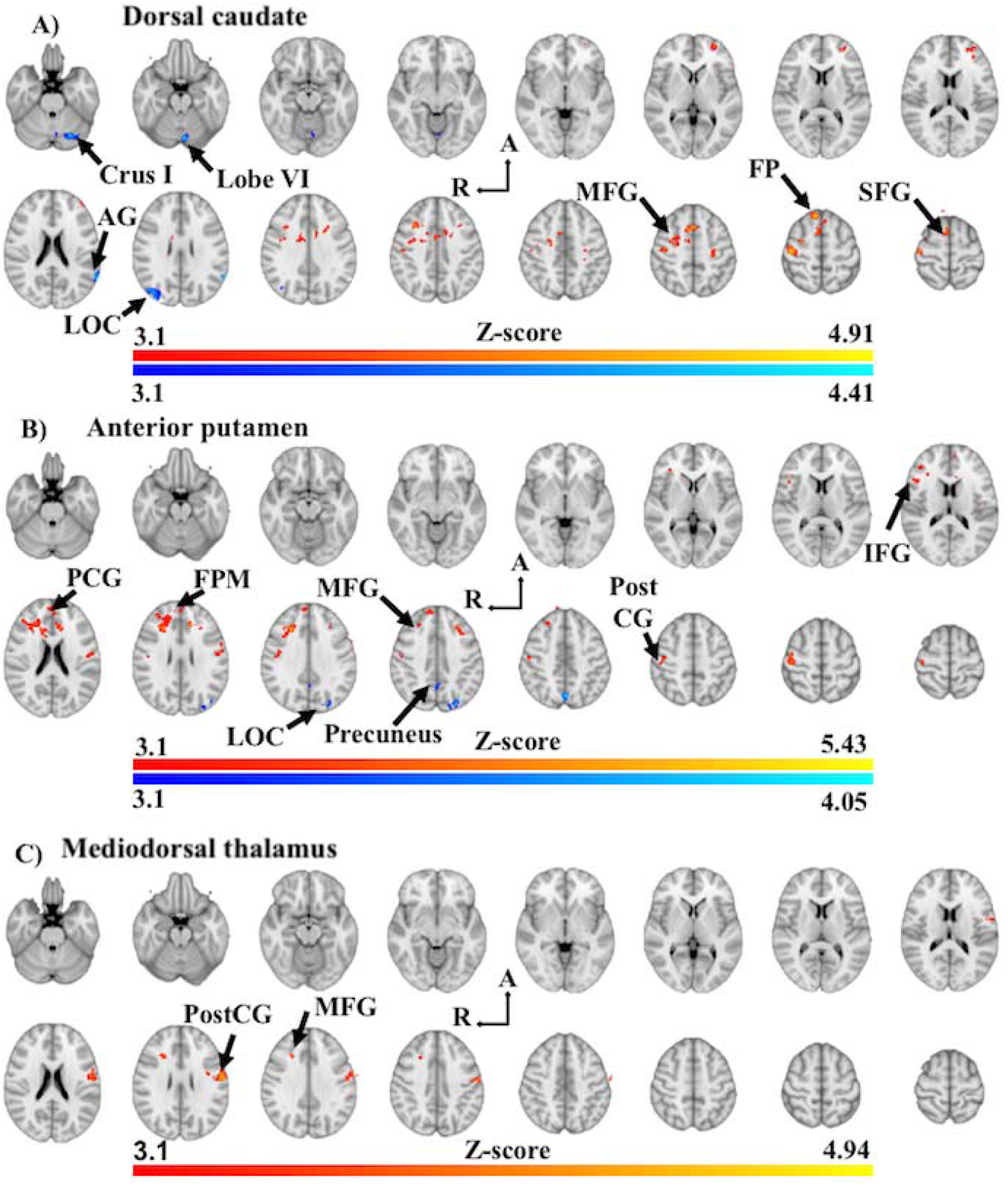
Significant between-group differences in functional connectivity of the dorsal caudate (A), anterior putamen (B) and mediodorsal thalamus (C) in PD-CU compared to Controls. Z-score statistic (Gaussianized) images thresholded non-parametrically using clusters determined by Z > 3.1 and a corrected cluster significance of p < 0.05, overlaid on MNI 152 T1 1mm standard axial brain image (Z = -26mm ascending to 64mm; 6mm interslice distance). Warm colours represent increased in functional connectivity in PD-CU, while shades of blue represent decreased functional connectivity in PD-CU. Abbreviations: Crus I, Crus I of cerebellum; Lobe VI, posterior cerebellar lobule VI; AG, angular gyrus; LOC, lateral occipital gyrus; A, anterior; R, right hemisphere; MFG, middle frontal gyrus; FP, frontal pole; SFG, superior frontal gyrus; PCG, paracingulate gyrus; FPM, frontal pole (medial); PostCG, postcentral gyrus.

##### Anterior putamen

Analysis of the anterior putamen in PD-CU compared to Controls found clusters of increased FC at the bilateral middle frontal gyri (BA8, 9), left frontal pole (BA10), bilateral postcentral gyri (BA2, 3), left paracingulate gyrus (BA32) and right inferior frontal gyrus (BA44) (Figure 2B; Table 2). These clusters extended to include local maxima at the bilateral anterior cingulate cortex (BA24), right frontal pole (BA10), left superior frontal gyrus (BA9), bilateral precentral gyri (BA4, 6) and bilateral central operculum (BA42) (Figure 2B; Supplementary Information Table 2). We also found clusters of decreased FC at the left lateral occipital cortex (BA7) and right precuneus (BA7) (Figure 2B; Table 2). These clusters extended to include a local maxima at the left precuneus (Figure 2B; Supplementary Information Table 2).

##### Mediodorsal thalamus

Analysis of the mediodorsal thalamus in PD-CU compared to Controls found clusters of increased FC at the left postcentral gyrus (BA1) and the right middle frontal gyrus (BA8) (Figure 2C; Table 2). These clusters extended to include local maxima at the left postcentral gyrus (BA3) and the left precentral gyrus (BA4, 6) (Figure 2C; Supplementary Information Table 3). No significant clusters of decreased FC were found for the mediodorsal thalamus.

**Table 3:** Functional connectivity differences in PD-MCI compared to PD-CU.

| Seed region | # | FC | Brain region | Voxels | Z | MNI |  |  |
| --- | --- | --- | --- | --- | --- | --- | --- | --- |
|  |  |  |  |  |  | x | y | z |
| Dorsal caudate | | ⇔ | - | - | - | - | - | - |
| Anterior putamen | | ⇔ | - | - | - | - | - | - |
| Mediodorsal thalamus | 23 | $\Downarrow$ | R paracingulate (BA32) | 137 | 3.92 | 2 | 46 | 30 |

**Key:** Coordinates of peak Z-score voxel location for areas of significant functional connectivity differences in PD-MCI compared to PD-CU (family-wise error corrected $p < 0.05$ ). Anatomical locations are based on the Harvard-Oxford Cortical and Subcortical Atlases in FSLEyes.
Abbreviations: #, cluster number; FC, functional connectivity; $\Downarrow$ , decrease in functional connectivity; ⇔ , no significant functional connectivity changes; R, right hemisphere; BA, Brodmann Area; Z, maximum Z-value for cluster; MNI, coordinates for location of maximum Z-value, defined in the Montreal Neurological Institute T1 standard space.

#### 3.2.2 PD-MCI compared to PD-CU

##### Mediodorsal thalamus

Analysis of the mediodorsal thalamus in PD-MCI compared to PD-CU found one significant cluster of decreased FC at the right paracingulate gyrus (BA32) (Figure 3; Table 3). This cluster extended to include local maxima at the medial frontal gyrus (BA9) and right frontal pole (BA8) (Figure 4; Supplementary Information Table 4). No significant clusters of increased FC were found. There were no significant FC differences to the dorsal caudate or anterior putamen when comparing PD-MCI and PD-CU.

**Table 4:** Functional connectivity differences in PDD compared to PD-MCI.

| Seed region | # | FC | Brain region | Voxels | Z | MNI |  |  |
| --- | --- | --- | --- | --- | --- | --- | --- | --- |
|  |  |  |  |  |  | x | y | z |
| Dorsal caudate | | ⇔ | - | - | - | - | - | - |
| Anterior putamen | | ⇔ | - | - | - | - | - | - |
| Mediodorsal thalamus | 24 | $\Downarrow$ | R posterior cingulate cortex (BA23) | 106 | 4.07 | 6 | -38 | 26 |

**Key:** Coordinates of peak Z-score voxel location for areas of significant functional connectivity differences in PDD compared to PD-MCI (family-wise error corrected $p < 0.05$ ). Anatomical locations are based on the Harvard-Oxford Cortical and Subcortical Atlases in FSLEyes. Abbreviations: #, cluster number; FC, functional connectivity; $\Downarrow$ , decrease in functional connectivity; ⇔ , no significant functional connectivity changes; R, right hemisphere; BA, Brodmann Area; Z, maximum Z-value for cluster; MNI, coordinates for location of maximum Z-value, defined in the Montreal Neurological Institute T1 standard space.

**Figure 3:**
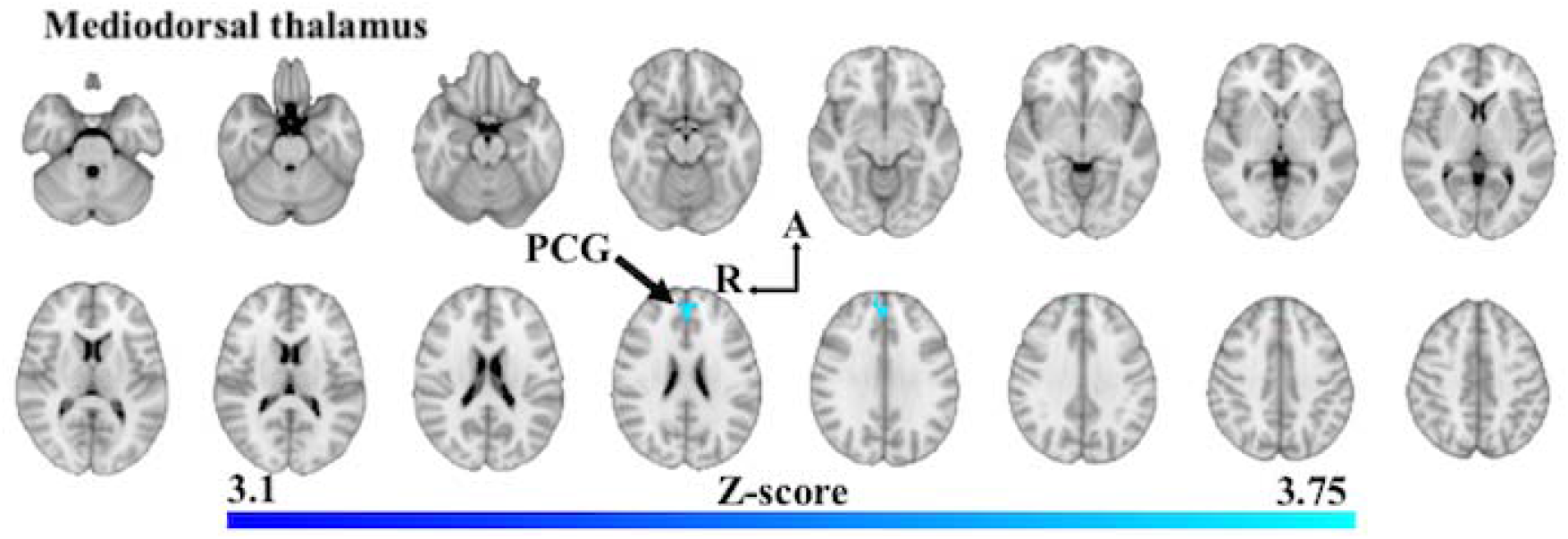
Significant between-group differences in functional connectivity of the mediodorsal thalamus in PD-MCI compared to PD-CU. Z-score statistic (Gaussianized) images thresholded non-parametrically using clusters determined by Z > 3.1 and a corrected cluster significance of *p* < 0.05, overlaid on MNI 152 T1 1mm axial standard brain image (Z = -30.5mm ascending to 45.5mm; 5mm interslice distance). Shades of blue show areas of decreased in functional connectivity in PD-MCI. Abbreviations: A, anterior; R, right hemisphere; PCG, paracingulate gyrus.

**Figure 4:**
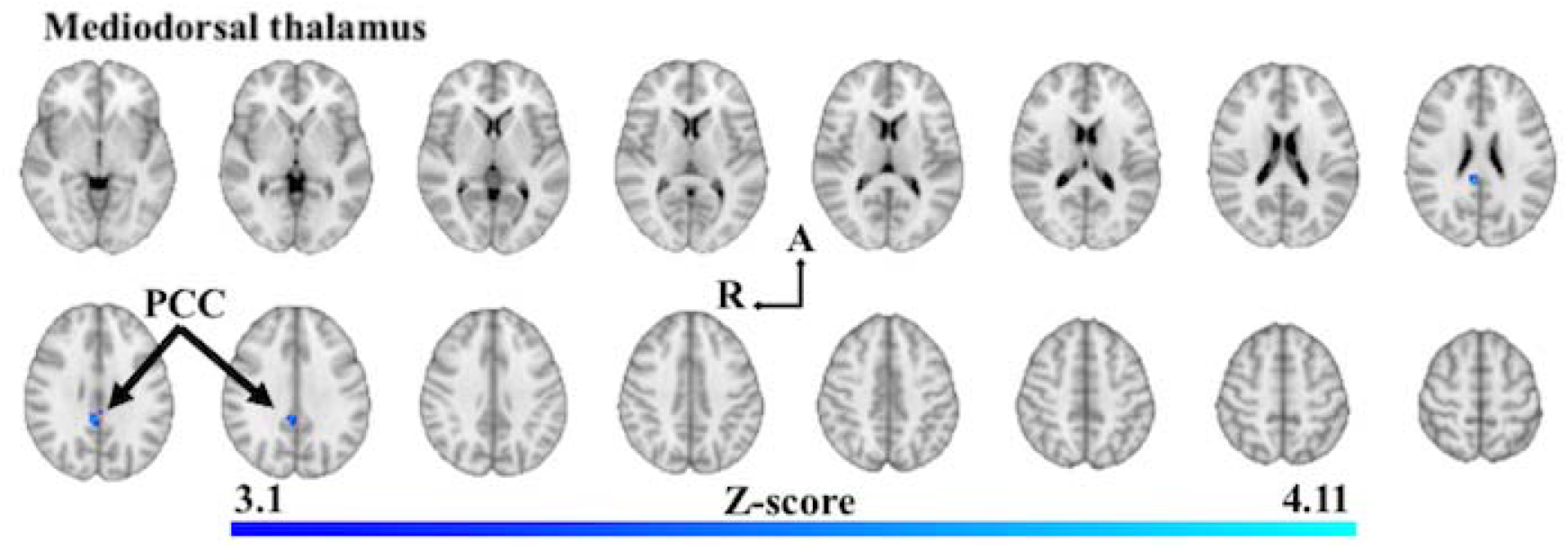
Significant between-group differences in functional connectivity of the mediodorsal thalamus in PDD compared to PD-MCI. Z-score statistic (Gaussianized) images thresholded non-parametrically using clusters determined by Z > 3.1 and a corrected cluster significance of *p* < 0.05, overlaid on MNI 152 T1 1mm axial standard brain image (Z = -3mm ascending to 57mm; 4mm interslice distance). Shades of blue show areas of decreased in functional connectivity in PDD. Abbreviations: PCC, Posterior cingulate cortex; R, right hemisphere; A, anterior.

#### 3.2.3 PDD compared to PD-CU

There were no significant FC differences to the dorsal caudate, anterior putamen or mediodorsal thalamus when comparing PD-CU subjects to PDD subjects.

#### 3.2.4. PDD compared to PD-MCI

##### Mediodorsal thalamus

Analysis of the mediodorsal thalamus in PDD compared to PD-MCI found one cluster of decreased FC at the right posterior cingulate cortex (BA23) (Figure 4; Table 4; Supplementary Information Table 5). There were no significant FC differences of the dorsal caudate or anterior putamen when comparing PDD with PD-MCI.

### 3.3 Volumetric analyses

Pairwise comparisons of volumes (Supplementary Information Table 6 and 7) revealed the following significant differences between experimental groups: the left caudate was significantly reduced in PDD compared to PD-CU (*p* = 0.009) and Controls (*p* < 0.001) with a large effect size (η^2^ = 0.177) (Figure 6A). The right caudate was significantly reduced in PDD compared to PD-CU (*p* = 0.021) and Controls (*p* = 0.001) also with a large effect size (η^2^ = 0.143) (Figure 5A). The left putamen was significantly reduced in PDD compared to PD-CU (*p* = 0.005) and Controls (*p* = 0.05) with a large effect size (η^2^ = 0.193) (Figure 5B). The right putamen was significantly reduced in PDD compared to Controls (*p* = 0.016) with a medium effect size (η^2^ = 0.107) (Figure 6B). The left thalamus was significantly reduced in PDD compared to PD-CU (*p* = 0.011) and Controls (*p* = 0.037) with a medium effect size (η^2^ = 0.108) (Figure 5C). The right thalamus was significantly reduced in PDD compared to PD-CU (*p* = 0.002) with a medium effect size (η^2^ = 0.136) (Figure 6C).

**Figure 5:**
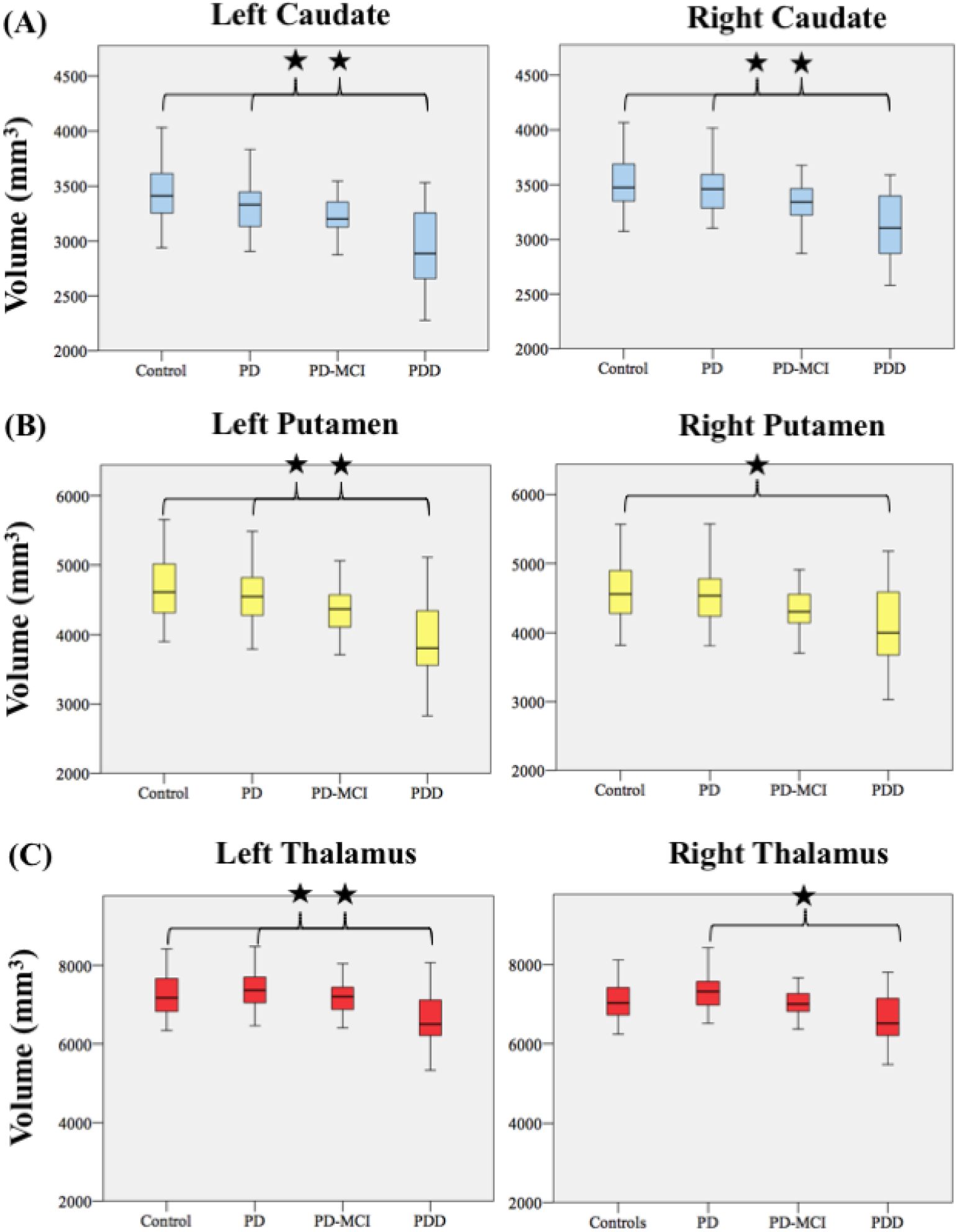
Pairwise comparison of volumes between PD subgroups and Controls. (A) Caudate (B) putamen and (C) thalamus. Volumes presented are estimated marginal mean values for each experimental group derived from MANCOVA models controlling for age and ICV. *, indicates a pairwise comparison with a significant *p*-value <0.05 confidence interval correction: Bonferroni.

**Figure 6:**
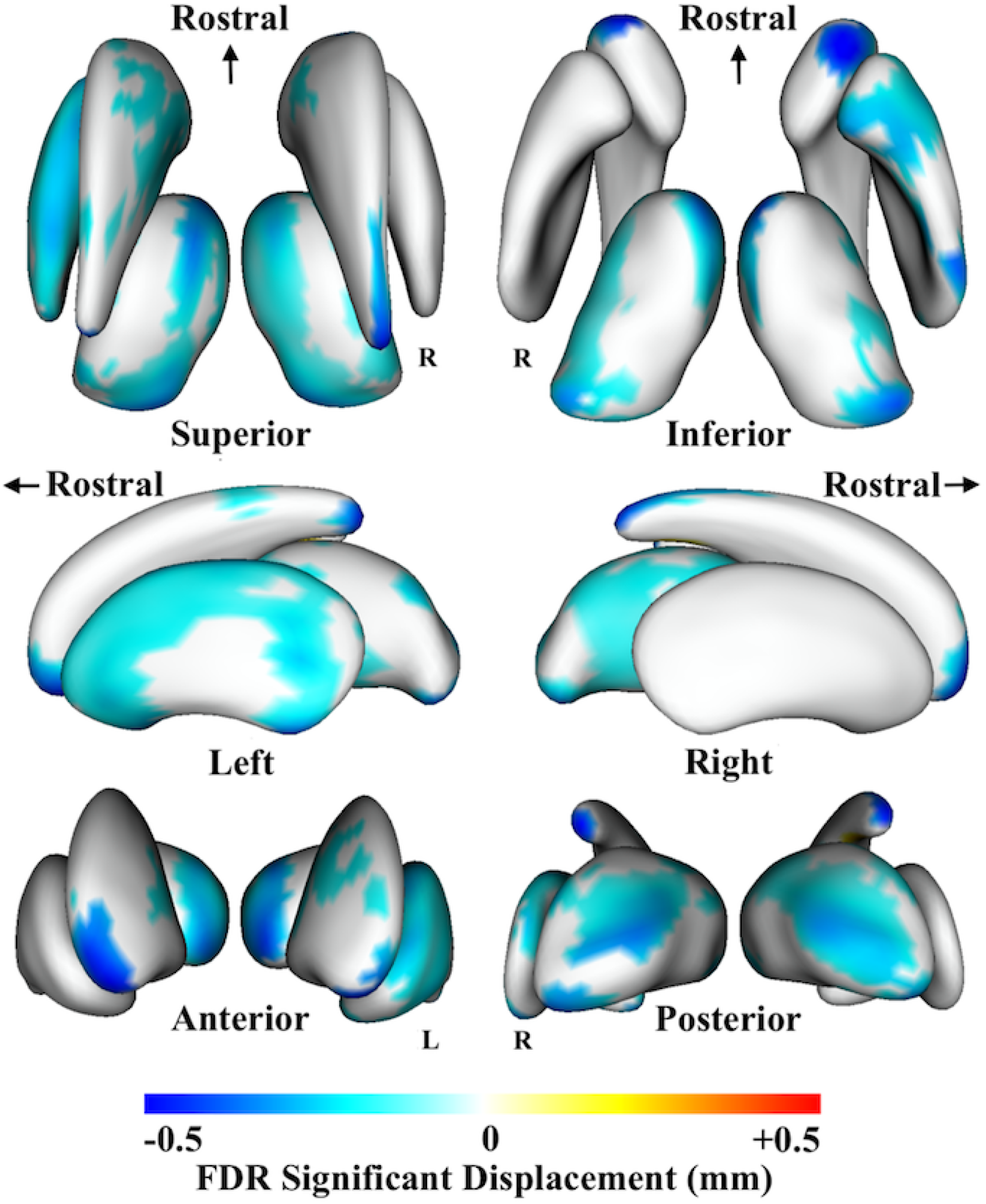
SPHARM-PDM shape analysis of the caudate nucleus, putamen and thalamus in PDD compared to PD-CU subjects. Neuroanatomical surface areas of FDR-corrected mean difference displacement maps. Displacement colour scale corresponds to the millimetres of deflation/inflation of the surface in that region in mm; warmer colours correspond to surface expansion in PDD while cooler colours correspond to surface deflation in PDD. The bilateral thalami sit medially, the caudate nuclei rostrally and the putamen are the most lateral of the three structures. Abbreviations: R, right hemisphere; FDR, false-discovery rate; Left, looking at the structures from the left side of the brain; R, looking at the structures from the right side of the brain.

### 3.4 SPHARM-PDM shape analyses

SPHARM-PDM shape analysis of the caudate, putamen and thalamus found widespread deflation to the surface of these structures in PDD compared to PD-CU subjects. Surface deflation can be observed at the bilateral rostral as well as dorsomedial surfaces areas of the caudate (Figure 6). Widespread deflation was also found on the surface of the left putamen, at the anterior, dorsal and posterior regions. Significant and widespread deflation was found across the medial surface of the bilateral thalami, potentially corresponding to surface regions of the anterior and mediodorsal thalamic nuclei. Surface deflation was also found across the posterior-ventral surfaces of the thalami, potentially corresponding to the pulvinar as well as medial and lateral geniculate bodies. No FDR-corrected significant shape changes were found when comparing PD-CU to PD-MCI or PD-MCI to PDD.

## 4. Discussion

This study investigates caudate nucleus, putamen and thalamus functional connectivity (FC) and morphology alterations associated with cognitive impairment and dementia in PD. Our study contributes new knowledge by showing how important functional subdivisions of these structures are impacted across cognitive disease stages.

Our findings of increased FC between the dorsal caudate, anterior putamen and mediodorsal thalamus with the pre- and postcentral gyri support a recent meta-analysis indicating that PD is associated with increased FC of these cortical regions (Ji et al., 2018). After weighing the relative contributions of seed-ROIs, this meta-analysis demonstrated that seeds placed within the caudate, putamen and thalamus contributed most to this result. Our findings also support work demonstrating increases in FC between a basal ganglia-thalamic network and areas of the prefrontal cortex and anterior/paracingulate gyri (Gorges et al., 2015). This study argues that such hyper-connectivity in cognitively unimpaired PD subjects may be representative of the recruitment of additional resources in the brain to maintain cognitive performance (Gorges et al., 2015). A common response to neurological disruption is hyper-connectivity of brain circuits, and it is suggested that such a response may reflect compensatory mechanisms in the brain to maintain normal levels of neuronal functioning (Hillary et al., 2015). Our results provide some support for this hypothesis and may signal how resources can be recruited to maintain ‘normal’ cognitive functioning. These increases in FC between the dorsal caudate and regions across the prefrontal cortex and anterior/paracingulate gyri may relate to changes within the executive control network (ECN). This network is comprised of dorsal and ventrolateral prefrontal, frontoinsular, lateral parietal, middle prefrontal cortices, anterior and paracingulate gyri, as well as subcortical sites at the mediodorsal and anterior thalamic nuclei and the dorsal caudate (Beckmann et al., 2005, Seeley et al., 2007). The ECN has been validated on a large scale and has shown that activity within this network corresponds strongly with several paradigms of cognition in task-based FC analyses (Smith et al., 2009).

Our data also indicate that PD-CU is associated with a reduction in FC between the dorsal caudate and the cerebellum. This result supports previous work demonstrated decreases in FC between the caudate nucleus and an ‘extended brainstem’ region encompassing the cerebellum (Hacker et al., 2012). These findings are significant because loss of FC between the caudate and the cerebellum may serve as a viable candidate for PD related gait difficulties, postural instability and freezing, which can be linked to dysfunction in cerebellar circuitry (Hacker et al., 2012). Our data also indicate that PD-CU subjects have decreased FC between the dorsal caudate/anterior putamen and the precuneus and angular gyrus. This finding supports the work demonstrating decreases in FC between nodes in the DMN, including lateral and medial parietal regions (Amboni et al., 2015).

While our results support a number of previous research findings, they contrast with similar work, necessitating explication. One group has demonstrated no significant differences in FC of the anterior striatum in a PD cohort (when placed on medication) compared to Control subjects (Bell et al., 2015), while also finding no differences in FC of seed-ROIs placed in the thalamus and brain regions within intrinsic connectivity networks. Similarly, our data contrast with work showing decreased FC between the caudate and putamen and brain regions across the frontal lobe, as well as *hypo*-connectivity of the thalamus with insular cortices (Agosta et al., 2014). There are a number of methodological considerations that may explain these discrepancies. The former work involved a younger PD cohort to the current study, which brings into question the impact of age related changes in FC (Fair et al., 2008), while the latter work focussed on a PD cohort with unilateral disease presentation, studying FC of the ‘more affected’ and ‘less affected’ hemispheres (Agosta et al., 2014). This approach increases the specificity of their analysis, however it makes comparisons with the present data difficult as we averaged FC signals from seeds within both hemispheres at each timepoint.

PD-MCI subjects showed decreased FC of the mediodorsal thalamus with the paracingulate gyrus, medial frontal gyrus and frontal pole compared to cognitively unimpaired PD subjects. Our findings of decreased FC of the mediodorsal thalamus support previous work indicating that PD-MCI is associated with lower FC between a basal ganglia-thalamic intrinsic connectivity network and the anterior/paracingulate gyri compared to a cognitively unimpaired PD cohort (Gorges et al., 2015). Decreased FC between the mediodorsal thalamus and medial prefrontal brain regions provides evidence for the important link between midline nodes within the ECN and cognitive function. As outlined, structures within this network are related to cognitive function (Beckmann et al., 2005, Seeley et al., 2007), as they work together to provide bias signals to other areas of the brain to implement and maintain cognitive control processes (Miller and Cohen, 2001). While we have demonstrated increased FC of nodes within this network in PD-CU subjects, here we demonstrate that PD-MCI subjects have a reduced FC between the mediodorsal thalamus and paracingulate cortices. While traditionally thought to be subserve affective functions, the cingulate has emerged as an important structure associated with higher cognitive processes, and has been shown to be particularly vulnerable to disease processes where executive control is impaired (Carter et al., 2000). Stronger FC between nodes of the ECN has been shown to be positively correlated with superior executive task performance of subjects (Seeley et al., 2007). If the corollary holds true, poorer cognitive performance, as our PD-MCI subjects demonstrate, might be associated with reduced FC within this network, which we have demonstrated.

PDD subjects showed decreased FC of the mediodorsal thalamus with the posterior posterior cingulate cortex (PCC) compared to PD-MCI subjects, supporting the findings from graph theoretical research (Zhan et al., 2018). Decreases in FC between these regions is significant due to the crucial role played by the PCC in cognitive functioning as well as the role of the mediodorsal thalamus and PCC within the default-mode network (DMN). The functional role played by the PCC involves elements of memory consolidation, attention and the control required to balance internally and externally focussed thoughts (Leech and Sharp, 2014). The PCC is affected in neurodegenerative diseases like Alzheimer’s disease (Buckner et al., 2009, Greicius et al., 2004, Mevel et al., 2011) and atrophy of the PCC has been demonstrated in PD subjects with dementia (Melzer et al., 2012). The PCC has important structural and also functional connectivity with the thalamus (Cunningham et al., 2017) and it has been suggested that the structure may play an important role balancing the interaction between brain regions in the prefrontal cortex that are involved cognitive processes and posterior nodes within the DMN (Greicius et al., 2003). These researchers suggested that the thalamus is uniquely positioned to act as an intermediary structure in the brain that balances the interaction between these regions due to its extensive cortico-cortical connectivity (Greicius et al., 2003). Our research supports this idea, highlighting how the FC between the mediodorsal thalamus and the PCC is impacted in PD subjects with dementia.

Our research found no evidence of morphological alterations to the caudate, putamen or thalamus in PD-CU subjects compared to Controls, supporting previous studies (Lee et al., 2014, Mak et al., 2014, Mckeown et al., 2008, Menke et al., 2014, Messina et al., 2011, Nemmi et al., 2015, Tinaz et al., 2011). We also found no evidence of volume loss to the caudate, putamen or thalamus in PD-MCI compared to Controls, supporting a previous study (Amboni et al., 2015). Volumetric changes in PDD were found in all three structures, supporting previous work (Burton et al., 2004, Melzer et al., 2012, Summerfield et al., 2005). Shape analysis demonstrated widespread surface deflation in PDD compared to PD-CU subjects, fine-tuning these volumetric results. Regarding the caudate, surface deflation was found primarily at the rostral and dorsomedial surfaces, areas with putative connectivity to the executive regions of the frontal lobe as well as the rostral motor regions (Tziortzi et al., 2014). These regions of the caudate are also known to demonstrate dopaminergic depletion (Kish et al., 1988). Surface deflation was found at the anterior, dorsal and posterior putamen surface regions areas with putative connectivity to the executive regions of the frontal lobe as well as caudal motor regions (Tziortzi et al., 2014). Widespread surface deflation was found primarily across the medial surface of the bilateral thalami, corresponding to surface regions of the anterior and mediodorsal thalamic nuclei with connections to the dorsolateral and prefrontal cortices (Behrens et al., 2003). Supporting this finding, this region of the thalamus displays significant cell loss in PD patients post-mortem (Halliday, 2009). Our findings support a previous study that found no significant difference between PD-MCI and cognitively unimpaired PD subjects using a vertex-wise shape analysis method (Mak et al., 2014). Our morphological data on PD-CU subjects are also consistent with previous studies which indicate that PD is not associated with localized surface alterations (Lee et al., 2014, Menke et al., 2014, Messina et al., 2011) however they contrast with the results of two groups who found localised surface deflation in PD (Garg et al., 2015, Mckeown et al., 2008). As raised previously, methodological inconsistencies between studies may be a significant factor behind these differences.

This research was performed in a cross-sectional manner, logically assuming a sequential temporal relationship between PD-CU, PD-MCI and PDD groupings. Categorisation of PD subjects in this way is a potential limitation, as we cannot be sure that all participants with PD-CU will progress to PD-MCI, then to PDD. While there is strong evidence that PD-MCI does represent an intermediate cognitive disease stage between cognitively unimpaired PD and PDD (Litvan et al., 2012), future research should follow a longitudinal design to better study how functional connectivity and morphology of basal ganglia-thalamocortical circuits are related to cognitive impairment in PD. This would have the added benefit of reducing the influence of the significant disease heterogeneity in PD which has been shown to have a significant impact on the reproducibility of results in this field of research (Badea et al., 2017).

This study demonstrated how the functional connectivity of subcortical hubs within basal ganglia-thalamocortical circuits are implicated in PD between cognitive disease stages. Functional connectivity changes are found in cognitively unimpaired PD subjects, who display no morphological changes, while PD-MCI subjects and PDD subjects both display decreases in functional connectivity of the mediodorsal thalamus with the anterior and posterior cingulate cortices respectively. Our research highlights the importance of the caudate, putamen and thalamus as core structures within key intrinsic connectivity networks in the brain, and indicates that a breakdown in functional connectivity of the mediodorsal thalamus is tied to mild cognitive impairment and dementia in PD.

## Data Availability

Anonymized data will be shared by request from any qualified investigator for the sole purpose of replicating procedures and results presented in the article and as long as data transfer is in agreement with EU legislation on the general data protection regulation. Data underlying the results described in our manuscript are available from at Attention: Conor Owens-Walton Academic Unit of Psychiatry & Addiction Medicine Australian National University Medical School Building 4, Level 2, Canberra Hospital Woden, A.C.T. 2605 AUSTRALIA or

## Acknowledgement

The authors are indebted to all patients and control subjects who participated in this study. CO-W would like to acknowledge the Australian National University for their funding support via the University Research Scholarship. This project is an initiative of the Swedish BioFINDER Study, of whom DvW and OH are steering committee members, and also the AUSSIE network coordinated by JCLL at the Australian National University Medical School, who self-funds related expenses. DJ reports no funding sources. MW has received consulting fees and honoraria from Actelion Pharmaceuticals and receives royalties from ACER (Australian Council of Educational research) for the NUCOG, a pencil and paper cognitive assessment tool (not used in this study. Work at the SH, DvW and OH research center was supported by the European Research Council, the Swedish Research Council, the Knut and Alice Wallenberg foundation, the Marianne and Marcus Wallenberg foundation, the Strategic Research Area MultiPark (Multidisciplinary Research in Parkinson’s disease) at Lund University, the Swedish Alzheimer Foundation, the Swedish Brain Foundation, The Parkinson foundation of Sweden, The Parkinson Research Foundation, the Skåne University Hospital Foundation, and the Swedish federal government under the ALF agreement. OH has acquired research support (for the institution) from Roche, GE Healthcare, Biogen, AVID Radiopharmaceuticals and Euroimmun. In the past 2 years, he has received consultancy/speaker fees (paid to the institution) from Biogen and Roche. The authors declare no competing interests. Funding sources had no role in the design and conduct of the study, in the collection, analysis, interpretation of the data or in the preparation, review or approval of the manuscript.

## Supplementary Information

*Supplementary_information_final.docx*

## Footnotes

1 Abbreviations used in this research paper: PD, Parkinson disease; PD-CU, cognitively unimpaired Parkinson disease; MCI, mild cognitive impairment; PD-MCI, Parkinson disease patients with mild cognitive impairment; PDD, Parkinson disease patients with dementia; FC, functional connectivity; UPDRS-III, ‘Unified Parkinson’s Disease Rating Scale Part-III’ test; MMSE, ‘Mini Mental State Examination’; ROI, region-of-interest; GLM, general linear model; LEDD, levodopa-equivalent daily dosage; SPHARM-PDM, spherical harmonic parametrization three-dimensional point distribution model; FDR; false-discovery rate; ECN, executive control network; PDD, posterior cingulate cortex; DMN; default-mode network.

## References

Agosta, F., Caso, F., Stankovic, I., Inuggi, A., Petrovic, I., Svetel, M., Kostic, V. S. & Filippi, M. 2014. Cortico-striatal-thalamic network functional connectivity in hemiparkinsonism. Neurobiology of Aging, 35, 2592–2602.

Amboni, M., Tessitore, A., Esposito, F., Santangelo, G., Picillo, M., Vitale, C., Giordano, A., Erro, R., De Micco, R. & Corbo, D. 2015. Resting-state functional connectivity associated with mild cognitive impairment in Parkinson’s disease. Journal of neurology, 262, 425–434.

Andersson, J. L. R., Jenkinson, M. & Smith, S. M. 2007a. Non-linear optimisation. FMRIB technical report TR07JA1.

Andersson, J. L. R., Jenkinson, M. & Smith, S. M. 2007b. Non-linear optimisation, aka spatial normalisation. FMRIB technical report TR07JA2.

Badea, L., Onu, M., Wu, T., Roceanu, A. & Bajenaru, O. 2017. Exploring the reproducibility of functional connectivity alterations in Parkinson’s disease. PLoS ONE, 12, e0188196.

Beckmann, C. F., Deluca, M., Devlin, J. T. & Smith, S. M. 2005. Investigations into restingstate connectivity using independent component analysis. Philos Trans R Soc Lond B Biol Sci, 360, 1001–13.

Beckmann, C. F. & Smith, S. M. 2004. Probabilistic independent component analysis for functional magnetic resonance imaging. IEEE Trans Med Imaging, 23, 137–52.

Behrens, T. E., Johansen-Berg, H., Woolrich, M. W., Smith, S. M., Wheeler-Kingshott, C. A., Boulby, P. A., Barker, G. J., Sillery, E. L., Sheehan, K., Ciccarelli, O., Thompson, A. J., Brady, J. M. & Matthews, P. M. 2003. Non-invasive mapping of connections between human thalamus and cortex using diffusion imaging. Nature Neuroscience, 6, 750–7.

Bell, P. T., Gilat, M., O’callaghan, C., Copland, D. A., Frank, M. J., Lewis, S. J. G. & Shine, J. M. 2015. Dopaminergic basis for impairments in functional connectivity across subdivisions of the striatum in Parkinson’s disease. Human Brain Mapping, 36, 1278–1291.

Broeders, M., De Bie, R., Velseboer, D. C., Speelman, J. D., Muslimovic, D. & Schmand, B. 2013. Evolution of mild cognitive impairment in Parkinson disease. Neurology, 81, 346–352.

Buckner, R. L., Sepulcre, J., Talukdar, T., Krienen, F. M., Liu, H., Hedden, T., Andrews-Hanna, J. R., Sperling, R. A. & Johnson, K. A. 2009. Cortical hubs revealed by intrinsic functional connectivity: mapping, assessment of stability, and relation to Alzheimer’s disease. Journal of Neuroscience, 29, 1860–1873.

Burton, E. J., Mckeith, I. G., Burn, D. J., Williams, E. D. & O’brien, J. T. 2004. Cerebral atrophy in Parkinson’s disease with and without dementia: a comparison with Alzheimer’s disease, dementia with Lewy bodies and controls.

Caballero-Gaudes, C. & Reynolds, R. C. 2017. Methods for cleaning the BOLD fMRI signal. Neuroimage, 154, 128–149.

Carter, C. S., Macdonald, A. M., Botvinick, M., Ross, L. L., Stenger, V. A., Noll, D. & Cohen, J. D. 2000. Parsing executive processes: strategic vs. evaluative functions of the anterior cingulate cortex. Proceedings of the National Academy of Sciences of the United States of America, 97, 1944–1948.

Chen, F. X., Kang, D. Z., Chen, F. Y., Liu, Y., Wu, G., Li, X., Yu, L. H., Lin, Y. X. & Lin, Z. Y. 2016. Gray matter atrophy associated with mild cognitive impairment in Parkinson’s disease. Neuroscience Letters, 617, 160–5.

Cohen, J. 1992. Statistical power analysis. Current directions in psychological science, 1, 98–101.

Cunningham, S. I., Tomasi, D. & Volkow, N. D. 2017. Structural and functional connectivity of the precuneus and thalamus to the default mode network. Hum Brain Mapp, 38, 938–956.

Damoiseaux, J. S., Rombouts, S. a. R. B., Barkhof, F., Scheltens, P., Stam, C. J., Smith, S. M. & Beckmann, C. F. 2006. Consistent resting-state networks across healthy subjects. Proceedings of the National Academy of Sciences of the United States of America,103, 13848-13853.

Emre, M., Aarsland, D., Brown, R., Burn, D. J., Duyckaerts, C., Mizuno, Y., Broe, G. A., Cummings, J., Dickson, D. W., Gauthier, S., Goldman, J., Goetz, C., Korczyn, A., Lees, A., Levy, R., Litvan, I., Mckeith, I., Olanow, W., Poewe, W., Quinn, N., Sampaio, C., Tolosa, E. & Dubois, B. 2007. Clinical diagnostic criteria for dementia associated with Parkinson’s disease. Movment Disorders, 22, 1689-707; quiz 1837.

Fahn, S. & Elton, R. (eds.) 1987. Members of the UPDRS development committee: The unified Parkison disease rating scale, Florham Park, NJ.: Macmillan Health Care Information.

Fair, D. A., Cohen, A. L., Dosenbach, N. U., Church, J. A., Miezin, F. M., Barch, D. M., Raichle, M. E., Petersen, S. E. & Schlaggar, B. L. 2008. The maturing architecture of the brain’s default network. Proceedings of the National Academy of Sciences, 105, 4028–4032.

Folstein, M. F., Folstein, S. E. & Mchugh, P. R. 1975. “Mini-mental state”: a practical method for grading the cognitive state of patients for the clinician. Journal of psychiatric research, 12, 189–198.

Garg, A., Appel-Cresswell, S., Popuri, K., Mckeown, M. J. & Beg, M. F. 2015. Morphological alterations in the caudate, putamen, pallidum, and thalamus in Parkinson’s disease. Frontiers in Neuroscience, 9.

Gelb, D. J., Oliver, E. & Gilman, S. 1999. Diagnostic criteria for Parkinson disease. Archives of neurology, 56, 33–39.

Genovese, C. R., Lazar, N. A. & Nichols, T. 2002. Thresholding of statistical maps in functional neuroimaging using the false discovery rate. Neuroimage, 15, 870–878.

Goldman, J. G., Holden, S. K., Litvan, I., Mckeith, I., Stebbins, G. T. & Taylor, J. P. 2018. Evolution of diagnostic criteria and assessments for Parkinson’s disease mild cognitive impairment. Movement Disorders, 33, 503–510.

Gorges, M., Muller, H. P., Lule, D., Pinkhardt, E. H., Ludolph, A. C. & Kassubek, J. 2015. To rise and to fall: functional connectivity in cognitively normal and cognitively impaired patients with Parkinson’s disease. Neurobiology of Aging, 36, 1727–1735.

Greicius, M. D., Krasnow, B., Reiss, A. L. & Menon, V. 2003. Functional connectivity in the resting brain: a network analysis of the default mode hypothesis. Proc Natl Acad Sci U S A, 100, 253–8.

Greicius, M. D., Srivastava, G., Reiss, A. L. & Menon, V. 2004. Default-mode network activity distinguishes Alzheimer’s disease from healthy aging: Evidence from functional MRI. Proceedings of the National Academy of Sciences of the United States of America, 101, 4637–4642.

Greve, D. N. & Fischl, B. 2009. Accurate and robust brain image alignment using boundary-based registration. Neuroimage, 48, 63–72.

Haber, S. N. 2003. The primate basal ganglia: parallel and integrative networks. Journal of chemical neuroanatomy, 26, 317–330.

Hacker, C. D., Perlmutter, J. S., Criswell, S. R., Ances, B. M. & Snyder, A. Z. 2012. Resting state functional connectivity of the striatum in Parkinson’s disease. Brain, 135, 3699–3711.

Halliday, G. M. 2009. Thalamic changes in Parkinson’s disease. Parkinsonism & Related Disorders, 15, Supplement 3, S152–S155.

Hillary, F. G., Roman, C. A., Venkatesan, U., Rajtmajer, S. M., Bajo, R. & Castellanos, N. D. 2015. Hyperconnectivity is a fundamental response to neurological disruption. Neuropsychology, 29, 59–75.

Hwang, K., Bertolero, M. A., Liu, W. B. & D’esposito, M. 2017. The human thalamus is an integrative hub for functional brain networks. Journal of Neuroscience, 37, 5594–5607.

Jenkinson, M., Bannister, P., Brady, M. & Smith, S. 2002. Improved optimization for the robust and accurate linear registration and motion correction of brain images. Neuroimage, 17, 825–41.

Jenkinson, M., Beckmann, C. F., Behrens, T. E. J., Woolrich, M. W. & Smith, S. M. 2012. FSL. Neuroimage, 62, 782–790.

Jenkinson, M. & Smith, S. 2001. A global optimisation method for robust affine registration of brain images. Medical image analysis, 5, 143–156.

Ji, G.-J., Hu, P., Liu, T.-T., Li, Y., Chen, X., Zhu, C., Tian, Y., Chen, X. & Wang, K. 2018. Functional Connectivity of the Corticobasal Ganglia–Thalamocortical Network in Parkinson Disease: A Systematic Review and Meta-Analysis with Cross-Validation. Radiology, 287, 172183.

Khan, A. R., Hiebert, N. M., Vo, A., Wang, B. T., Owen, A. M., Seergobin, K. N. & Macdonald, P. A. 2018. Biomarkers of Parkinson’s disease: Striatal sub-regional structural morphometry and diffusion MRI. Neuroimage: Clinical.

Kish, S. J., Shannak, K. & Hornykiewicz, O. 1988. Uneven pattern of dopamine loss in the striatum of patients with idiopathic Parkinson’s disease. Pathophysiologic and clinical implications. New England Journal of Medicine, 318, 876–80.

Lee, H. M., Kwon, K.-Y., Kim, M.-J., Jang, J.-W., Suh, S.-I., Koh, S.-B. & Kim, J. H. 2014. Subcortical grey matter changes in untreated, early stage Parkinson’s disease without dementia. Parkinsonism & related disorders, 20, 622–626.

Leech, R. & Sharp, D. J. 2014. The role of the posterior cingulate cortex in cognition and disease. Brain, 137, 12–32.

Litvan, I., Goldman, J. G., Troster, A. I., Schmand, B. A., Weintraub, D., Petersen, R. C., Mollenhauer, B., Adler, C. H., Marder, K., Williams-Gray, C. H., Aarsland, D., Kulisevsky, J., Rodriguez-Oroz, M. C., Burn, D. J., Barker, R. A. & Emre, M. 2012. Diagnostic criteria for mild cognitive impairment in Parkinson’s disease: Movement Disorder Society Task Force guidelines. Movment Disorders, 27, 349–56.

Looi, J. C. & Walterfang, M. 2013. Striatal morphology as a biomarker in neurodegenerative disease. Molecular Psychiatry, 18, 417–24.

Looi, J. C., Walterfang, M., Nilsson, C., Power, B. D., Van Westen, D., Velakoulis, D., Wahlund, L. O. & Thompson, P. M. 2014. The subcortical connectome: Hubs, spokes and the space between - a vision for further research in neurodegenerative disease. Australian and New Zealand Journal of Psychiatry.

Mak, E., Bergsland, N., Dwyer, M. G., Zivadinov, R. & Kandiah, N. 2014. Subcortical atrophy is associated with cognitive impairment in mild Parkinson disease: a combined investigation of volumetric changes, cortical thickness, and vertex-based shape analysis. American Journal of Neuroradiology, 35, 2257–64.

Mckeown, M. J., Uthama, A., Abugharbieh, R., Palmer, S., Lewis, M. & Huang, X. 2008. Shape (but not volume) changes in the thalami in Parkinson disease. BMC neurology, 8, 8.

Melzer, T. R., Watts, R., Macaskill, M. R., Pitcher, T. L., Livingston, L., Keenan, R. J., Dalrymple-Alford, J. C. & Anderson, T. J. 2012. Grey matter atrophy in cognitively impaired Parkinson’s disease. Journal of Neurology, Neurosurgery and Psychiatry, 83, 188.

Menke, R. A., Szewczyk□Krolikowski, K., Jbabdi, S., Jenkinson, M., Talbot, K., Mackay, C.E. & Hu, M. 2014. Comprehensive morphometry of subcortical grey matter structures in early□stage Parkinson’s disease. Human brain mapping, 35, 1681–1690.

Messina, D., Cerasa, A., Condino, F., Arabia, G., Novellino, F., Nicoletti, G., Salsone, M., Morelli, M., Lanza, P. L. & Quattrone, A. 2011. Patterns of brain atrophy in Parkinson’s disease, progressive supranuclear palsy and multiple system atrophy. Parkinsonism & related disorders, 17, 172–176.

Mevel, K., Chetelat, G., Eustache, F. & Desgranges, B. 2011. The default mode network in healthy aging and Alzheimer’s disease. International journal of Alzheimer’s disease, 2011, 535816.

Miller, E. K. & Cohen, J. D. 2001. An integrative theory of prefrontal cortex function. Annu Rev Neurosci, 24, 167–202.

Nemmi, F., Sabatini, U., Rascol, O. & Peran, P. 2015. Parkinson’s disease and local atrophy in subcortical nuclei: insight from shape analysis. Neurobiology of Aging, 36, 424–33.

Obeso, J. A., Rodriguez-Oroz, M. C., Rodriguez, M., Lanciego, J. L., Artieda, J., Gonzalo, N. & Olanow, C. W. 2000. Pathophysiology of the basal ganglia in Parkinson’s disease. Trends in neurosciences, 23, S8–S19.

Owens-Walton, C., Jakabek, D., Li, X., Wilkes, F. A., Walterfang, M., Velakoulis, D., Van Westen, D., Looi, J. C. L. & Hansson, O. 2018. Striatal changes in Parkinson disease: An investigation of morphology, functional connectivity and their relationship to clinical symptoms. Psychiatry Research: Neuroimaging, 275, 5–13.

Patenaude, B., Smith, S. M., Kennedy, D. N. & Jenkinson, M. 2011. A Bayesian model of shape and appearance for subcortical brain segmentation. Neuroimage, 56, 907–922.

Pitcher, T. L., Melzer, T. R., Macaskill, M. R., Graham, C. F., Livingston, L., Keenan, R. J., Watts, R., Dalrymple-Alford, J. C. & Anderson, T. J. 2012. Reduced striatal volumes in Parkinson’s disease: a magnetic resonance imaging study. Translational Neurodegeneration, 1, 68–76.

Poewe, W., Seppi, K., Tanner, C. M., Halliday, G. M., Brundin, P., Volkmann, J., Schrag, A.-E. & Lang, A. E. 2017. Parkinson disease. Nature Reviews Disease Primers, 3, 17013.

Power, B. D. & Looi, J. C. 2015. The thalamus as a putative biomarker in neurodegenerative disorders. Australian and New Zealand Journal of Psychiatry, 0004867415585857.

Salimi-Khorshidi, G., Douaud, G., Beckmann, C. F., Glasser, M. F., Griffanti, L. & Smith, S. M. 2014. Automatic denoising of functional MRI data: combining independent component analysis and hierarchical fusion of classifiers. Neuroimage, 90, 449–468.

Seeley, W. W., Menon, V., Schatzberg, A. F., Keller, J., Glover, G. H., Kenna, H., Reiss, A. L. & Greicius, M. D. 2007. Dissociable intrinsic connectivity networks for salience processing and executive control. J Neurosci, 27, 2349–56.

Smith, S. M. 2002. Fast robust automated brain extraction. Human brain mapping, 17, 143–155.

Smith, S. M., Fox, P. T., Miller, K. L., Glahn, D. C., Fox, P. M., Mackay, C. E., Filippini, N., Watkins, K. E., Toro, R., Laird, A. R. & Beckmann, C. F. 2009. Correspondence of the brain’s functional architecture during activation and rest. Proc Natl Acad Sci U S A, 106, 13040–5.

Sporns, O., Tononi, G. & Kötter, R. 2005. The Human Connectome: A Structural Description of the Human Brain. PLoS Comput Biol, 1.

Sterling, N. W., Du, G., Lewis, M. M., Dimaio, C., Kong, L., Eslinger, P. J., Styner, M. & Huang, X. 2013. Striatal shape in Parkinson’s disease. Neurobiology of Aging, 34, 2510–2516.

Strafella, A. P. 2013. Anatomical and functional connectivity as a tool to study brain networks in Parkinson’s disease. Movement Disorders, 28, 411–412.

Styner, M., Oguz, I., Xu, S., Brechbuhler, C., Pantazis, D., Levitt, J. J., Shenton, M. E. & Gerig, G. 2006. Framework for the Statistical Shape Analysis of Brain Structures using SPHARM-PDM. Insight J, 242–250.

Summerfield, C., Junque, C., Tolosa, E., Salgado-Pineda, P., Gomez-Anson, B., Marti, M. J., Pastor, P., Ramirez-Ruiz, B. & Mercader, J. 2005. Structural brain changes in Parkinson disease with dementia: a voxel-based morphometry study. Arch Neurol, 62, 281–5.

Tinaz, S., Courtney, M. G. & Stern, C. E. 2011. Focal cortical and subcortical atrophy in early Parkinson’s disease. Movement Disorders, 26, 436–441.

Tziortzi, A. C., Haber, S. N., Searle, G. E., Tsoumpas, C., Long, C. J., Shotbolt, P., Douaud, G., Jbabdi, S., Behrens, T. E. J., Rabiner, E. A., Jenkinson, M. & Gunn, R. N. 2014. Connectivity-Based Functional Analysis of Dopamine Release in the Striatum Using Diffusion-Weighted MRI and Positron Emission Tomography. Cerebral Cortex, 24, 1165–1177.

Williams-Gray, C. H., Foltynie, T., Brayne, C. E. G., Robbins, T. W. & Barker, R. A. 2007. Evolution of cognitive dysfunction in an incident Parkinson’s disease cohort. Brain, 130, 1787–1798.

Woolrich, M. W., Behrens, T. E. J., Beckmann, C. F., Jenkinson, M. & Smith, S. M. 2004. Multilevel linear modelling for FMRI group analysis using Bayesian inference. Neuroimage, 21, 1732–1747.

Woolrich, M. W., Ripley, B. D., Brady, M. & Smith, S. M. 2001. Temporal autocorrelation in univariate linear modeling of FMRI data. Neuroimage, 14, 1370–1386.

Worsley, K. 2001. Statistical analysis of activation images. Functional MRI: an introduction to methods, 14, 251–270.

Zhan, Z.-W., Lin, L.-Z., Yu, E.-H., Xin, J.-W., Lin, L., Lin, H.-L., Ye, Q.-Y., Chen, X.-C. & Pan, X.-D. 2018. Abnormal resting-state functional connectivity in posterior cingulate cortex of Parkinson’s disease with mild cognitive impairment and dementia. CNS Neuroscience & Therapeutics, 24, 897–905.

Zhang, Y., Brady, M. & Smith, S. 2001. Segmentation of brain MR images through a hidden Markov random field model and the expectation-maximization algorithm. IEEE Transactions on Medical Imaging, 20, 45–57.

